# Fetal exposure to paracetamol is associated with altered markers of ovarian development and reduced uterine volume in girls: the COPANA study

**DOI:** 10.64898/2026.04.23.26351576

**Authors:** MB Fischer, G Mola, K Sundberg, L Scheel, KB Wraae, AL Rom, H Frederiksen, RA Anderson, M Assens, AM Andersson, L Priskorn, JH Petersen, HK Hegaard, KM Main, DM Kristensen, A Juul, CP Hagen

## Abstract

**Study question:** is fetal exposure to paracetamol associated with markers of ovarian function in infancy?

**Summary answer:** Mild to moderate doses of prenatal paracetamol exposure, assessed by detailed maternal reports and urinary measurements, is associated with ovarian morphology and activity as well as reduced size of estrogen-responsive tissues in infant girls.

**What is known already:** Maternal use of paracetamol is widespread. Across multiple independent animal studies, fetal exposure consistently impairs the formation of primordial ovarian follicles, causing subfertility and premature estropause in female offspring.

**Study design, size, duration:** The Copenhagen Analgesic study (COPANA) is a single center, prospective, observational cohort study conducted at the Copenhagen University Hospital – Rigshospitalet, Denmark (March 2020 to November 2022).

**Participants/materials, setting, methods;:** *COPANA cohort:* 3425 eligible participants. In total, 685 healthy, singleton pregnant women of Caucasian origin were enrolled in the first trimester of pregnancy, 302 girls examined at follow up. Exclusion criteria: maternal diabetes or thyroid disease, pre- or post-term delivery, or severe infant illness. Exposure: pregnant women reported paracetamol use biweekly and provided first-trimester urinary samples which were analyzed for paracetamol levels (LC-MS/MS) and adjusted for urinary osmolarity (n = 299). Girls were classified by timing of exposure: early fetal life (<17 weeks, n = 92), mid-late fetal life (≥17 weeks, n = 67), or unexposed controls (n = 143). A subgroup of girls was exposed exclusively in early fetal life (n = 22).

*Independent confirmatory cohort:* 1210 girls followed from infancy to adolescence. Exposure: maternal self-reported *any* use of paracetamol during pregnancy reported in early third trimester (yes/no).

**Main results and the role of chance:** Early fetal exposure was associated with reduced ovarian volume (−0.11 cm^3^, 95% CI −0.19 to −0.03) and uterine volume (−0.16 cm^3^, −0.32 to −0.01), whereas mid-late fetal exposure was associated with fewer ovarian follicles (−1.05, −1.71 to −0.39) compared to unexposed girls. AMH levels were lower in girls exposed exclusively in early fetal life (−0.45 SDS, −0.87 to −0.03) compared to unexposed girls. Maternal urinary paracetamol concentrations were inversely associated with ovarian and uterine volume as well as breast tissue diameter. In an independent cohort, fetal paracetamol exposure was associated with reduced uterine volume at puberty (−4.11 cm^3^, −7.29 to −0.92) and smaller ovarian volume in adolescence (−2.76 cm^3^, −4.82 to −0.70).

**Limitations, reasons for caution:** The design of the study allows evaluation of exposure- outcome associations, while causality is strengthened by experimental models demonstrating comparable effects. Residual confounding by indication cannot be completely excluded, although results were robust after accounting for fever and other maternal factors. The analytic design of the current study limited our ability to evaluate whether frequency or patterns of paracetamol use influenced the observed associations.

**Wider implications of the findings:** The consistency of findings across different assessment measures and cohorts, in combination with parallel evidence from animal studies, suggests potential long-term implications for female reproductive health.

**Study funding/competing interest(s):** This research was supported by Rigshospitalets Research Council under grant (E-22717-21), Læge Sofus Carl Emil Friis og hustru Doris Friis’ Legat (F-23936-01), Aase og Ejnar Danielsens Foundation (20-10-0367), Helsefonden (20-B-0388), Axel Muusfeldt Foundation (2020-0385) and The Danish Centre for Endocrine Disrupting Substances (CeHoS) (2022-23219). The authors have nothing to declare.

**Trial registration number:** ClinicalTrials.gov ID: NCT0436922

## 1. Introduction

The foundation of adult reproductive function is established in utero through the development and differentiation of the fetal gonads. Females are born with a defined number of primordial follicles that deplete throughout their reproductive lifespan inevitably leading to menopause (Baker, 1963; Wallace and Kelsey, 2010). Fetal establishment of the primordial follicle pool is therefore crucial for female reproductive health.

In early fetal life, oogenesis involves rapid germ cell proliferation and the formation of primordial follicles in gestational weeks 10-17 (Baker, 1963; Forabosco and Sforza, 2007). The transition from mitosis to meiosis and subsequent arrest in prophase I is critical for oocyte survival and establishing the follicle pool (Hartshorne *et al*., 2009).

From mid- to late fetal life, mitosis declines as more germ cells enter meiosis and folliculogenesis begins (Forabosco and Sforza, 2007; Kurilo, 1981). These sequential developmental phases represent highly sensitive windows in which adverse exposure may have long-term consequences for female reproductive health.

Minipuberty is a transient activation of the hypothalamic-pituitary-gonadal (HPG) axis during infancy resulting in increased ovarian activity including ovarian follicle growth and feedback mechanisms resembling those of adult women (Fischer, Mola, Rom, *et al*., 2024; Ljubicic *et al*., 2022). This provides a unique opportunity to assess ovarian activity in early infancy.

Paracetamol (*N*-acetyl-*p*-aminophenol, otherwise known as paracetamol) is sold over the counter, reducing its perception as a pharmacologic agent with potential side effects (Bauer *et al*., 2021). Due to its analgesic and antipyretic properties, it is used by more than 50% of women during pregnancy, and it passes the placenta freely (Nitsche *et al*., 2017).

In rodent studies, fetal paracetamol exposure reduces ovarian follicle reserve by up to 40% in female offspring, leading to impaired fertility and shortened reproductive lifespan (Aleixo *et al*., 2020; Dean *et al*., 2016; Holm *et al*., 2016; Hurtado-Gonzalez *et al*., 2018; Johansson *et al*., 2016; Rossitto *et al*., 2019; Wu *et al*., 2024).

Comparable findings have been observed with acetylsalicylic acid and other non-steroidal anti-inflammatory drugs, likely due to their irreversible inhibition of prostaglandin synthases (Kristensen *et al*., 2012; Mazaud-Guittot *et al*., 2013). Acetylsalicylic acid is commonly prescribed during pregnancy to women at risk of complications (Roberge *et al*., 2018).

Beyond pharmaceutical use, paracetamol exposure may also occur environmentally through aniline, an industrial precursor found in textiles and pesticides, which is metabolized to paracetamol in the liver (Kristensen *et al*., 2016; Pot E *et al*., 2022). Urinary paracetamol levels (U-paracetamol) provide an objective biomarker of recent exposure and complement self-reported medication.

While experimental and animal data consistently demonstrate the harmful effects of fetal exposure to paracetamol on gonadal development, evidence in humans remains limited. The Copenhagen Analgesic Study – COPANA is the first human study primarily designed to evaluate if fetal exposure to paracetamol is associated with impaired gonadal function. We assessed ovarian and uterine morphology as well as circulating levels of reproductive hormones in 302 infant girls during minipuberty. Findings were further evaluated in an independent confirmatory cohort, The Copenhagen Mother-Child Cohort, followed from fetal life through adolescence (Assens *et al*., 2020; Chellakooty *et al*., 2003; Hagen *et al*., 2015).

## 2. Methods

### 2.1 Population

The Copenhagen Analgesic study (COPANA) is a single center, prospective, observational cohort study conducted at the Copenhagen University Hospital - Rigshospitalet, Denmark (March 2020 to November 2022) (Fischer, Mola, Scheel, *et al*., 2024).

A total of 3,425 healthy pregnant women (Caucasian, BMI 18–35 kg/m^2^, no pre-existing diabetes or thyroid disease) were invited to participate after completing a routine web-based questionnaire (Q1) providing the obstetrics department with patient-reported health information. From these, 685 women were enrolled. To obtain a homogeneous study population, and to avoid conditions possibly affecting fetal development and hormone levels, women were excluded due to multiple pregnancy, gestational diabetes, thyroid disease, pre- or post-term delivery (< 37 GW or > 42 GW), and serious illness in the infant such as genetic conditions and birth defects. Women reporting substance use (other than nicotine or alcohol) were also excluded. Further, at inclusion, all women were asked to complete a questionnaire on general and reproductive health (Q2).

Participating families visited three times during the study; at the first trimester, at a third trimester fetal ultrasound scan, and for an infant examination approximately 3 months post-partum during minipuberty (n = 302 girls). For the study flowchart and timeline, see Supplementary Fig. S1.

### 2.2 Exposure to paracetamol

The Q1 questionnaire from which eligible women were invited was completed in mean (±SD) GW 9.8 (2.4) and entailed information about current use of mild analgesics (“Do you currently take any mild analgesic?” (Consumption: yes/no, rarely/1-2 times weekly/daily, drug name). From the time of inclusion, GW 14.1 (1.8), and throughout the study period, the pregnant women completed detailed electronic questionnaires (Q3) every two weeks concerning medicine consumption with a focus on mild analgesics (consumption: yes/no, drug name, dose pr. use (mg), indication). The first questionnaire on medicine consumption at inclusion covered the period from the invitation to inclusion. Detailed information on medicine intake is therefore available from completion of Q1 at GW 9.8 (2.4). The specific generic and brand names of all available mild analgesic products were listed in the questionnaire. After birth, the questionnaire included questions about medication given to the infant.

Two fetal exposure periods were predefined reflecting different essential aspects of follicle formation and thus potentially different vulnerability regarding exposure:

Exposure in early fetal life: maternal use of paracetamol from invitation to GA 17 weeks. This period is characterized by intense mitotic proliferation of the oogonia, followed by initiation of the first meiotic division.

Exposure in mid-late fetal life: maternal use of paracetamol in GA ≥ 17 weeks, a period where mitotic activity subsides, paralleled by an increasing rate of meiotic entry. Moreover, folliculogenesis is ongoing.

We grouped the girls according to initiation of exposure within the predefined exposure periods. Girls who were not exposed to paracetamol during fetal life were classified as “unexposed controls”. A subgroup of girls was exposed to paracetamol exclusively in early fetal life. Girls initially exposed in mid-late fetal life were, by definition, exclusively exposed in this period. In addition to self-reported paracetamol use, the pregnant women provided a urine sample at the inclusion visit, which was analyzed for urinary levels of paracetamol.

### 2.3 Third trimester ultrasound scan

At GW 29-34, all included pregnant women were offered a study specific ultrasound scan (Voluson E10, GE HealthCare). Fetal anogenital distance (AGD) was measured in the axial plane from the center of the anus to the posterior converge of the fourchette by a board-certified technician (LS) with at least three repeated measurements wherefrom the average was calculated.

### 2.4 Infant examination

All infants were examined during minipuberty (age mean (±SD) 106 days (13)) with weight and length measurements. AGDaf (ano-fourchettal) and AGDac (ano-clitoral) were measured using the TIDES method(Sathyanarayana *et al*., 2015). Breast tissue was assessed by palpation, and the diameter was measured with a ruler.

A transabdominal ultrasound scan was performed by a single experienced clinician (MBF), blinded to exposure status (Voluson E10, GE HealthCare). Girls were scanned with a full bladder in a supine position with a slight incline. One or both ovaries were identified in 203 girls (bilaterally in 136 girls and unilaterally in 67 girls (left: n = 22to right: n = 45)). The length (L) and width (W) were measured, and ovarian volume was calculated (L*W*W*π/6) separately for the left and right side, whereafter an average of the two measures was used when both ovaries were identified. Antral follicles larger than 2mm were counted. The sum of follicles from both ovaries was used as the outcome.

Left and right-side ovarian volume (r = 0.48, p < 0.001, n = 136, Spearmańs rho) and follicle count (r = 0.38, p < 0.001, n = 136) correlated positively. Therefore, in cases where only one ovary was visualized, we used that ovarian volume or follicle count (doubled) as representative data for the contralateral ovary.

The uterus was scanned in the sagittal view, where the uterine length (SAG, including the cervix) was measured and at the widest point, the anteroposterior width (AP) and the thickness of the endometrium. In the transverse view, the widest transverse diameter (TRV) of the uterine fundus was measured. Uterine volume was calculated (SAG*AP*TRV*π/6).

### 2.5 Serum and urine samples

Concentrations of reproductive hormones were measured from blood samples drawn from an antecubital vein (n=269/302, 89 %). Samples were clotted, centrifuged, and serum was stored at −20^◦^C until analysis. The peptide hormones, Anti-Müllerian hormone (AMH), follicle stimulating hormone (FSH), luteinizing hormone (LH) and inhibin B were analyzed using immunoassays. The concentration of estradiol was analyzed by isotope-diluted online-TurboFlow-liquid chromatography mass spectrometry (LC-MS/MS) (Frederiksen, Johannsen, *et al*., 2020). For details on assays, detection limits (LOD) and prioritization of analyses, see Supplementary Table S1. Maternal urine samples (n = 299/302 (99%)) were analyzed for the total (free and conjugated) concentration of paracetamol by isotope diluted LC-MS/MS with prior enzymatic deconjugation and adjusted for urinary dilution by osmolality adjustments, according to previous description (Frederiksen, Nielsen, *et al*., 2020). LOD for paracetamol was 0.48 µg/L.

### 2.6 Outcomes

Primary outcomes were predefined as markers of ovarian function: ovarian volume, number of follicles and circulating levels of AMH (Fischer, Mola, Scheel, *et al*., 2024). In the present cohort, uterine size and glandular breast tissue diameter are closely correlated to ovarian activity and estradiol levels at minipuberty(Fischer, Mola, Rom, *et al*., 2024), and we therefore added those as primary outcomes.

Secondary outcomes were defined as key parameters relating to the primary outcomes, e.g. AGD, endometrial thickness, as well as circulating levels of FSH, LH and inhibin B.

### 2.7 Statistics

Continuous data were summarized using mean (±SD) or median (IQR) as appropriate and compared between exposure groups using Student’s t-test or Mann-Whitney U-test, while categorical data were summarized as frequency (%) and compared using the chi-square test or Fisher’s exact test when expected cell counts were ≤ 5. To adjust infant hormone concentrations for sex and age at measurement, we calculated sex- and age-specific standard deviation scores (SDS). We used reference curves from a previous extensive longitudinal study(Busch *et al*., 2021) which were created using the generalized additive model for location, scale, and shape (GAMLSS) including the GAMLSS R package (Rigby and Stasinopoulos, 2014). Limited serum volumes occasionally prevented full hormone analysis. Values below the limit of detection (LOD) were assigned as LOD/2. LH was above LOD in just 47 % of cases and therefore dichotomized to above LOD (yes/no).

We performed linear regression analyses to estimate regression coefficients (β) and 95% confidence intervals (CI) for the association between pre-specified exposure windows (early/mid-late) and predefined outcomes (dependent variables) compared to unexposed controls. To assess a possible dose-response trend, accumulated maternal self-reported doses of paracetamol use during pregnancy, as well as urinary paracetamol concentrations, were evaluated by generating binary exposure variables at percentile thresholds (at 1 % increments) ranging from the 50th to the 95th percentile. For each cutoff, participants above the threshold were compared with those below using linear regression models. Plots of residuals and QQ plots were used to assess model assumptions. Because residual diagnostics indicated that one variable (ovarian volume) did not fully meet the assumptions of normality and homoscedasticity, all regression analyses were conducted using robust variance estimators, which leave point estimates unchanged but provide heteroskedasticity-consistent standard errors, yielding more reliable confidence intervals and significance tests. This approach also allowed the outcome variables to be analyzed on their original scales without transformation, facilitating the interpretation of regression coefficients in their natural units.

Regression analyses were performed unadjusted as well as adjusted. Covariates were selected a priori based on biological plausibility and prior experimental evidence (maternal use of acetylsalicylic acid (Mazaud-Guittot *et al*., 2013) and other non-steroidal anti-inflammatory drugs (Leverrier-Penna *et al*., 2018) and infant treatment with paracetamol(A. N. Ndeke, 2011), maternal self-reported diagnosis of polycystic ovarian syndrome(Barrett *et al*., 2018), maternal consumption of alcohol in the first trimester) or observed differences between exposure groups (maternal pre-pregnancy BMI, maternal use of nicotine products in the first trimester, conception by IVF). Moreover, fetal AGD was adjusted for gestational age at the time of ultrasound scan, and measurements of infant AGD, uterine sizes and breast tissue diameter was adjusted for infant age at the time of examination.

Using multiple linear regression, we estimated the independent effect of exposure in each fetal period on the outcome variable while accounting for potential overlap between exposure periods. For each exposure period, a “dummy variable” was created, representing any paracetamol use during that specific period (yes/no), regardless of exposure in other fetal periods. The regression model included both dummy variables (early/mid-late fetal exposure) as independent variables simultaneously.

To assess the robustness of the observed associations, we conducted a series of sensitivity analyses. Specifically, we repeated the univariate regression models while sequentially excluding infants based on the presence of each binary (yes/no) covariate at a time.

To assess the degree of confounding by indication, we conducted an additional univariate analysis excluding infants from mothers who reported fever as the indication for paracetamol use during pregnancy.

A two-sided p-value was considered statistically significant at p ≤ 0.05. Trends were noted for p-values between 0.05 and 0.1. All statistical analyses were performed using IBM SPSS Statistics 22.0 (IBM Corp. Released 2013. IBM SPSS Statistics for Windows, Version 22.0. Armonk, NY: IBM Corp., n.d.), and RStudio (RStudio Team (version 2024.04.2). RStudio: Integrated Development for R. RStudio, PBC, Boston, MA, USA).

### 2.8 Ethics

The COPANA Study (ClinicalTrials.gov ID: NCT04369222) was approved by the ethical committee of the Capital Region of Denmark (H-19044036) and the Danish Data Protection Agency (P-2019-330). Participants gave written informed consent. For infant participants, consent was obtained from all adults with legal guardianship.

## 3. Independent confirmatory cohort

The Copenhagen Mother-Child Cohort is a population-based, longitudinal, prospective pregnancy and birth cohort initiated in 1996 with inclusion of pregnant women from three university hospitals in the greater Copenhagen area. Only pregnant women with Danish-born and −raised parents and grandparents were included. In total, 1,210 girls born between 1997 and 2002 were included at birth (Chellakooty *et al*., 2003).

### 3.1 Exposure

While this cohort was not primarily designed to evaluate associations between gonadal function and fetal exposure to mild analgesics, information on maternal use of medication during pregnancy was collected by questionnaire at the beginning of the third trimester. Mothers were asked if they had used *any form* of medication during pregnancy (yes/no/do not know), and paracetamol was specifically listed as an example. We grouped girls according to exposure to paracetamol and/or NSAIDs at any point during pregnancy (n = 179) vs. unexposed controls (including “do not know” (n = 1,012)).

### 3.2 Examinations

Girls were examined at infancy: n = 1,083, age mean (±SD) 101 (24) days, puberty: n = 563, age 10.7 (1.5) years and adolescence: n = 317, age 16.05 (0.89) years. At all examinations, girls underwent blood sampling and at puberty and adolescence, pubertal development was assessed by Tanner staging (Marshall and Tanner, 1969).

At puberty, a subset of girls (n=121) had a magnetic resonance imaging (MRI) of the uterus and ovaries. Magnetic resonance imaging was performed with a 3-Tesla MRI (Magnetom Verioto Siemens AG) as previously described (Hagen *et al*., 2015). At adolescence, a transabdominal ultrasound scan of the uterus and ovaries (n = 317) was performed (Voluson E8 Ultrasound System (GE Healthcare Medical Systems, Zipf, Austria)) (Assens *et al*., 2020). Adolescent girls were examined during cycle day 2-5 when possible. For both MRI and TAUS, the uterus was measured in three planesto sagittal (SAG), width (AP) and transverse (TRV) and the volume was calculated assuming an ellipsoid shape (SAG*AP*TRV*π/6). Likewise, ovarian volume was calculated using the length, width and transverse length. The number of follicles was counted in each ovary, and the total follicle count was calculated as the sum of follicles from both ovaries.

### 3.3 Serum samples

Serum FSH and LH were measured in infancy by Delfia (Wallac, Inc, Turku, Finland, LODs = 0.06 and 0.05 IU/liter, respectively)(Chellakooty *et al*., 2003), and in puberty and adolescence by Delfia (PerkinElmer, Boston, MA., LOD = 0.05 IU/L) (Assens *et al*., 2020; Hagen *et al*., 2015). Serum inhibin B was measured in infancy by an enzyme-linked immunosorbent assay (ELISA, Oxford Bio-Innovation, LOD = 20 pg/ml)(Chellakooty *et al*., 2003), and in puberty and adolescence by the Beckman Coulter GenII assay (Beckman Coulter, Inc. Brea, CA., LOD = 3.0 pg/mL).

In infancy and puberty, AMH was measured using the Beckman Coulter enzyme immunometric assay generation I (IOT, Immunotech, Beckman Coulter) with a detection limit of 2.0 pmol/L. During the follow-up of adolescent girls, the Access immunoassay substituted the Immunometric assay generation I when this was no longer commercially available. The Access immunoassay has a detection limit of 0.14 pmol/L. Comparative studies between the two assays revealed negligible differences (1.3%), and no adjustments of results have therefore been performed.

### 3.4 Statistics

We performed linear regression analyses to estimate regression coefficients (β) and 95% confidence intervals (CI) for the association between serum sample analyses as well as uterine and ovarian morphology (puberty and adolescence) (dependent variables) and exposure compared to unexposed controls (yes/no) (independent variable). Pubertal outcomes were adjusted for tanner stage at time of the examination, and for adolescent outcomes all girls using hormonal contraceptives were excluded. To ensure comparability with the COPANA cohort, we additionally adjusted for gestational week at birth and maternal thyroid disease or diabetes at all examination points (infancy/puberty/adolescence).

The Copenhagen Mother-Child cohort was conducted according to the Helsinki II declaration and approved by the local Ethical Committee [reference no. (KF) 01-030/97] and the Danish Registry Agency (journal no. 1997-1200-074).

## 4. Results from the COPANA cohort

In total, use of paracetamol during pregnancy was reported by 159 women (53%) to initially exposed in early fetal life (n = 92, dose: median (IQR) 2.0 g (1.0 to 3.7)), initially in mid-late fetal life (n = 67, dose: 2.0 g (1.0 to 3.5)) and a sub-group of girls exclusively exposed in early fetal life (n = 22, dose: 1.0 g (1.0 to 2.6)). The remaining 143 girls were unexposed controls. The indications of use were headache or migraine (70 %), pain (predominantly from the musculoskeletal system (18 %), general sickness or cold (6 %), fever (5 %), and other reasons (1 %).

In total, 21 women were treated with acetylsalicylic acid for prevention of obstetrical complications from early- and throughout pregnancy. Other non-steroidal, anti-inflammatory drugs were used by 12 women. Post-partum, 33 infants were treated with paracetamol.

Overall, the mean (±SD) maternal age at birth was 33 (4) years, and the majority of pregnant women were nulliparous (76 %). There were no differences in birth weight or gestational age at birth between infants exposed during defined fetal periods and unexposed controls (Table 1). Summaries of predefined outcomes stratified by exposure groups are presented in Table 2.

**Table 1.** Maternal and infant characteristics stratified by exposure initially in early (< GA 17 weeks) or mid-late (GA ≥ 17 weeks) fetal life compared to unexposed controls.

|  | Total (n = 302) |  | Unexposed (n = 143) |  | Initially exposed in early fetal life (n = 92) |  | Initially exposed in mid-late fetal life (n = 67) |  |
| --- | --- | --- | --- | --- | --- | --- | --- | --- |
|  | N | Mean (±SD) / median (IQR) / n (%) | N | Mean (±SD) / median (IQR) / n (%) | N | Mean (±SD) / median (IQR) / n (%) | N | Mean (±SD) / median (IQR) / n (%) |
| **MOTHER** |  |  |  |  |  |  |  |  |
| Maternal age at delivery (years) | 302 | 33 (4) | 143 | 33 (4) | 92 | 34 (4) | 67 | 33 (4) |
| Maternal prepregnancy bmi (kg/m<sup>2</sup>) | 294 | 22.3 (2.7) | 138 | 22.2 (2.3) | 89 | **22.8 (3.1)\*** | 67 | 22.2 (2.3) |
| Nulliparous (yes) | 301 | 228 (76) | 142 | 110 (78) | 92 | 65 (71) | 67 | 54 (81) |
| Use of nicotin products in first trimester (yes) | 302 | 9.0 (3) | 143 | 2 (1) | 92 | **7 (8)\*\*** | 67 | 0 (0) |
| Alcohol consumption in first trimester (yes) | 301 | 102 (33.8) | 143 | 42 (29) | 91 | 35 (39) | 67 | 25 (37) |
| Polycystic ovarian syndrome (yes) | 298 | 22 (7.4) | 140 | 8 (6) | 91 | 9 (10) | 67 | 5 (8) |
| Mode of conception |  |  |  |  |  |  |  |  |
| IUI | 301 | 18 (6) | 143 | 8 (6) | 91 | 4 (4) | 67 | 6 (9) |
| IVF | 301 | 16 (5) | 143 | 4 (3) | 91 | **8 (9)\*** | 67 | 4 (6) |
| ICSI | 301 | 15 (5) | 143 | 6 (4) | 91 | 7 (8) | 67 | 2 (3) |
| Dose of paracetamol (g) | 159 | 3.0 (1.0 to 8.5) | 143 | - | 92 | 2.0 (1.0 to 3.7) | 67 | 2.0 (1.0 to 3.5) |
| **INFANT** |  |  |  |  |  |  |  |  |
| Birthweight (g) | 301 | 3489 (440) | 142 | 3512 (412) | 92 | 3473 (507) | 67 | 3461 (402) |
| Birthweight (SDS) | 301 | -0.46 (0.99) | 142 | -0.38 (0.96) | 92 | -0.50 (1.2) | 67 | -0.56 (0.84) |
| Gestational age at birth (days) | 302 | 282 (9) | 143 | 282 (10) | 92 | 282 (8) | 67 | 283 (9) |

GA: gestational age, BMI: body-mass index, IUI: intrauterine insemination, IVF: in vitro fertilization, ICSI: intracytoplasmic sperm injection, SDS: standard deviation score. P-values by Student's t test or Pearson Chi-square test, with Fisher's exact test applied when expected counts ≤ 5. \*Indicate associations with a p-value 0.05 ≤ 0.1 and
\*\*indicate a p-value ≤ 0.05.

**Table 2.** Summaries of predefined primary and secondary outcomes in girls exposed initially during early fetal life (GA < 17 weeks, n = 92), and mid-late fetal life (GA ≥ 17 weeks, n = 67) compared to unexposed girls (n = 143)

|  | Total (n = 302) |  | Unexposed (n = 143) |  | Initially exposed in early fetal life (n = 92) |  | Initially exposed in mid-late fetal life (n = 67) |  |
| --- | --- | --- | --- | --- | --- | --- | --- | --- |
|  | N | Mean (±SD) / median (IQR) / n (%) | N | Mean (±SD) / median (IQR) / n (%) | N | Mean (±SD) / median (IQR) / n (%) | N | Mean (±SD) / median (IQR) / n (%) |
| Uterine volume (cm<sup>3</sup>) | 230 | 1.28 (0.50) | 107 | 1.34 (0.60) | 73 | **1.18 (0.36)\*\*** | 50 | 1.30 (0.42) |
| Endometrial thickness (mm) | 255 | 0.24 (0.06) | 118 | 0.24 (0.07) | 80 | 0.23 (0.06) | 57 | 0.24 (0.06) |
| Ovarian volume (cm<sup>3</sup>) | 203 | 0.26 (0.17 to 0.45) | 98 | 0.27 (0.20 to 0.42) | 68 | 0.26 (0.17 to 0.45) | 37 | 0.28 (0.13 to 0.45) |
| Sum of total follicles | 203 | 4.39 (2.10) | 98 | 2.08 (0.50) | 68 | 2.07 (0.49) | 37 | **1.83 (0.48)\*\*** |
| Breast tissue diameter (cm) | 300 | 0.87 (0.51) | 142 | 0.89 (0.49) | 91 | 0.88 (0.51) | 67 | 0.81 (0.54) |
| Fetal AGD (cm) | 295 | 1.28 (0.23) | 139 | 1.27 (0.22) | 90 | 1.27 (0.23) | 66 | 1.30 (0.24) |
| AGDaf (cm) | 301 | 1.58 (0.33) | 142 | 1.56 (0.33) | 92 | 1.62 (0.32) | 67 | 1.57 (0.34) |
| AGDac (cm) | 301 | 3.67 (0.48) | 142 | 3.69 (0.46) | 92 | 3.69 (0.51) | 67 | 3.58 (0.47) |
| AMH (SDS) | 262 | -0.12 (1.04) | 121 | -0.14 (1.01) | 81 | -0.05 (1.13) | 60 | -0.20 (0.98) |
| Inhibin B (SDS) | 256 | 0.03 (0.87) | 118 | 0.02 (0.90) | 78 | 0.16 (0.82) | 60 | -0.12 (0.87) |
| FSH (SDS) | 255 | -0.06 (0.88) | 117 | -0.07 (0.92) | 79 | -0.08 (0.79) | 59 | -0.02 (0.93) |
| LH measurable (yes) | 255 | 112 (44) | 117 | 50 (43) | 79 | 36 (46) | 59 | 26 (44) |
| Estradiol (SDS) | 269 | 0.03 (1.04) | 124 | 0.10 (1.05) | 84 | 0.02 (0.98) | 61 | -0.09 (1.10) |

GA: gestational age, SDS: standard deviation score, AGD: anogenital distance, AGDaf: anogenital distance (ano-fourchettal), AGDac: anogenital distance (ano-clitoral), LH: luteinizing hormone, FSH: follicle stimulating hormone, AMH: anti-Müllerian hormone. P-values for continuous variables were obtained from univariate linear regression and categorical variables from Pearson Chi square test. \*\*indicate a p-value ≤ 0.05.

### 4.1 Early fetal life (GA < 17 weeks)

In adjusted models, girls exposed initially during early fetal life had reduced ovarian volume: β (95% CI) −0.11 cm^3^ (−0.19 to −0.03), p = 0.01, corresponding to a −41 % change (−70 to −11) compared to unexposed girls. Similarly, uterine volume was reduced: −0.18 cm^3^ (−0.35 to −0.01) p = 0.04, corresponding to a −13 % change (−26.0 to −0.7) compared to unexposed girls (Table 3, Fig. 1). Exposure exclusively in early fetal life was associated with reduced ovarian volume: −0.10 cm^3^ (−0.19 to −0.01), p = 0.05, as well as lower circulating levels of AMH: −0.45 SDS (−0.87 to −0.03), p = 0.04 (Supplementary Table S2).

**Fig. 1.**
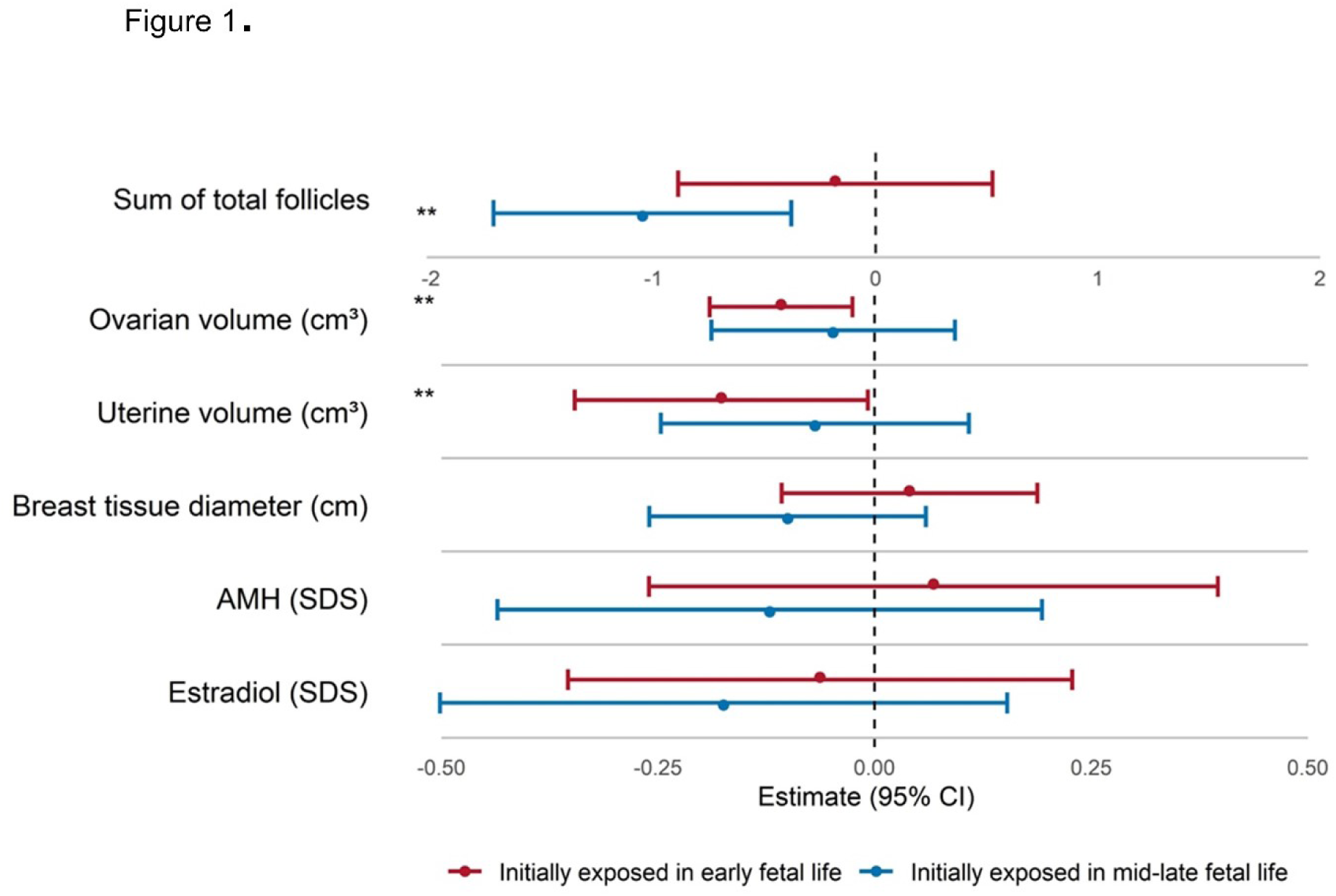
Forest plot of adjusted estimates of predefined primary outcomes by multiple linear regression models analyzing the association with exposure initially (n = 92) in early fetal life (GA < 17 weeks) and initially during mid-late fetal life (GA ≥ 17 weeks, n = 67) compared to unexposed controls (n = 143) by maternal self-reported use of paracetamol. Estimates are adjusted for maternal polycystic ovarian syndrome status, BMI before pregnancy, alcohol and nicotine use in the first trimester, conception by in vitro fertilization, maternal use of acetylic salicylic acid and other non-steroidal anti-inflammatory drugs, infant paracetamol treatment and infant age at examination (uterine volume, breast tissue diameter). SDS: standard deviation score, AMH: anti-Müllerian hormone. Dots represent the estimated effect size, and horizontal lines indicate 95% confidence intervals. *Indicates associations with a p-value of 0.05 ≤ 0.1 and **indicate a p-value ≤ 0.

**Table 3.** Unadjusted and adjusted associations between maternal self-reported use of paracetamol and predefined primary and secondary outcomes in girls exposed initially during early fetal life (GA < 17 weeks, n = 92) and mid-late fetal life (GA ≥ 17 weeks, n = 67), compared to unexposed girls (n = 143)

|  | Early fetal life |  | Mid-late fetal life |  |
| --- | --- | --- | --- | --- |
|  | Unadjusted | Adjusted | Unadjusted | Adjusted |
|  | Estimate (95% CI) | Estimate (95% CI) | Estimate (95% CI) | Estimate (95% CI) |
| Uterine volume (cm<sup>3</sup>) | **-0.16 (-0.3 to -0.02)\*\*** | **-0.18 (-0.35 to -0.01)\*\*** | -0.04 (-0.2 to 0.12) | -0.07 (-0.25 to 0.11) |
| Endometrial thickness (mm) | -0.02 (-0.2 to 0.15) | 0 (-0.02 to 0.02) | 0.01 (-0.19 to 0.2) | 0.01 (-0.01 to 0.03) |
| Ovarian volume (cm<sup>3</sup>) | -0.05 (-0.14 to 0.04) | **-0.11 (-0.19 to -0.03)\*\*** | -0.03 (-0.17 to 0.11) | -0.05 (-0.19 to 0.09) |
| Sum of total follicles | -0.06 (-0.72 to 0.6) | -0.18 (-0.88 to 0.52) | **-1.02 (-1.67 to -0.37)\*\*** | **-1.05 (-1.71 to -0.39)\*\*** |
| Breast tissue diameter (cm) | -0.01 (-0.14 to 0.12) | 0.04 (-0.11 to 0.19) | -0.08 (-0.23 to 0.07) | -0.1 (-0.26 to 0.06) |
| Fetal AGD (cm) | 0 (-0.06 to 0.06) | -0.03 (-0.09 to 0.03) | 0.02 (-0.04 to 0.09) | 0.01 (-0.05 to 0.07) |
| AGDaf (cm) | 0.06 (-0.03 to 0.14) | 0.01 (-0.12 to 0.14) | 0.01 (-0.09 to 0.11) | -0.1 (-0.24 to 0.04) |
| AGDac (cm) | 0 (-0.13 to 0.13) | 0.05 (-0.04 to 0.14) | **-0.11 (-0.25 to 0.02)\*** | -0.01 (-0.11 to 0.09) |
| AMH (SDS) | 0.09 (-0.21 to 0.39) | 0.07 (-0.26 to 0.4) | -0.06 (-0.37 to 0.24) | -0.12 (-0.43 to 0.19) |
| Inhibin B (SDS) | 0.14 (-0.11 to 0.38) | 0.18 (-0.07 to 0.43) | -0.14 (-0.41 to 0.13) | -0.14 (-0.42 to 0.14) |
| FSH (SDS) | -0.01 (-0.25 to 0.23) | 0.01 (-0.25 to 0.27) | 0.05 (-0.24 to 0.33) | 0.05 (-0.25 to 0.35) |
| Estradiol (SDS) | -0.08 (-0.36 to 0.2) | -0.06 (-0.35 to 0.23) | -0.2 (-0.53 to 0.13) | -0.17 (-0.49 to 0.15) |

Estimated mean differences with 95% confidence intervals (CI) from linear regression models, adjusted and unadjusted. Model adjusted for maternal polycystic ovarian syndrome status, BMI before pregnancy, alcohol and nicotine use in the first trimester, conception by in vitro fertilization, maternal use of acetylic salicylic acid and other non-steroidal anti-inflammatory drugs, infant paracetamol treatment and infant age at examination (uterine volume, AGDaf, AGDac, breast tissue diameter). Fetal AGD is moreover adjusted for GA at time of ultrasound scan. SDS: standard deviation score, AGD: anogenital distance, AGDaf: anogenital distance (ano-fourchettal), AGDac: anogenital distance (ano-clitoral), FSH: follicle stimulating hormone, AMH: anti-Müllerian hormone. \*indicate associations with a p-value $0.05 \leq 0.1$ and
\*\*indicate a p-value $\leq 0.05$ .

### 4.2 Mid-late fetal life (GA ≥ 17 weeks)

In adjusted models, girls exposed exclusively during mid-late fetal life had a reduced number of ovarian follicles: −1.05 (−1.71 to −0.39), p < 0.01 corresponding to −51 % (−82 to −19) compared to unexposed girls (Table 3, Fig.1).

Multiple regression analyses that simultaneously included both fetal exposure periods (early and mid-late), supported that the association between early fetal exposure to paracetamol and uterine volume was driven primarily by early fetal exposure, with no additional effect by exposure during mid-late fetal life. Similarly, reduced follicle count was associated with mid-late fetal exposure. In these analyses, we did not see a clear association between exposure periods and ovarian volume or circulating levels of reproductive hormones (Supplementary Table S3).

### 4.3 Dose-response trends

The total maternal paracetamol dose during pregnancy, based on self-reported data, was median (IQR) 3.0 g (1.0 to 8.5). Paracetamol was measurable in all urine samples from the mothers of included girls with a median (IQR) concentration of 64.5 ng/mL (34.6 to 134.9), (GW mean (±SD) 14.1 (1.8)). Both total dose of paracetamol and levels of maternal U-paracetamol were inversely associated with ovarian- and uterine volume. Moreover, levels of maternal U-paracetamol were inversely associated with breast tissue diameter (Fig.2). Percentile distribution of accumulated maternal self-reported doses of paracetamol use during pregnancy as well as first trimester urinary paracetamol concentrations are presented in Supplementary Table S4.

**Fig. 2.**
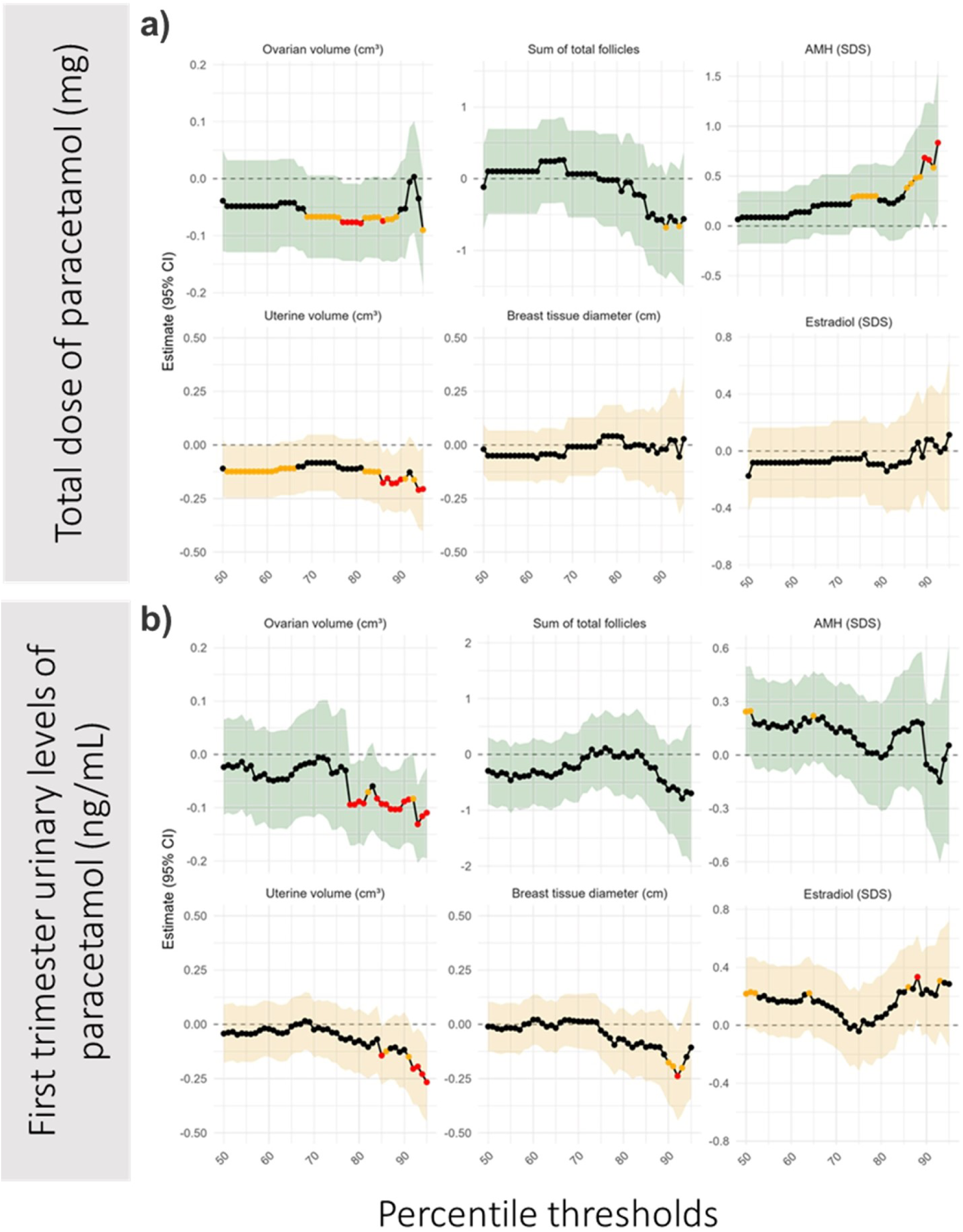
Dose-response relationships for (a) self-reported maternal paracetamol use and (b) urinary paracetamol concentrations in relation to predefined primary outcomes. Figure shows effect estimates (standardized β coefficients) across the percentile thresholds comparing participants above versus below the given paracetamol exposure threshold using linear regression models. Black lines indicate regression estimates, shaded areas represent 95% CIs. Points with p ≤ 0.05 are highlighted in red, and those with 0.05 < p ≤ 0.1 in orange. Models were adjusted for maternal polycystic ovary syndrome (PCOS) status, pre-pregnancy BMI, alcohol and nicotine use during the first trimester, conception by in vitro fertilization, maternal use of acetylsalicylic acid or other non-steroidal anti-inflammatory drugs, infant paracetamol treatment, and infant age at examination (for uterine volume and breast tissue diameter outcome). SDS: standard deviation score, AMH: anti-Müllerian hormone.

Overall, sensitivity analyses confirmed that associations between exposure to paracetamol in early, or mid-late fetal life and key outcomes were not affected by excluding infants based on covariate characteristics (data not shown) or the indication of maternal fever (Supplementary Table S5).

## 5. Independent confirmatory cohort

In total, use of paracetamol and/or non-steroidal anti-inflammatory drugs during pregnancy was reported by 179 women (18%). Mother and infant characteristics are listed in Supplementary Table S6 and descriptive summaries of outcomes in Supplementary Table S7.

At puberty, girls exposed to paracetamol and/or non-steroidal anti-inflammatory drugs during pregnancy had reduced uterine volume: −4.11 cm^3^ (−7.29 to −0.92), p = 0.01 as well as reduced levels of circulating inhibin B: −14.12 pg/mL (−25.20 to −3.03), p = 0.02 and FSH just reaching statistical significance: −0.61 IU/L (−1.21 to −0.00), p = 0.05. At adolescence, girls exposed to paracetamol and/or non-steroidal anti-inflammatory drugs during pregnancy had reduced ovarian volume: −2.76 cm^3^ (−4.82 to −0.70), p = 0.01 (Supplementary Table S8).

## 6. Discussion

To our knowledge, this is the first prospective human cohort study specifically designed to evaluate associations between fetal exposure to paracetamol and postnatal ovarian function. We demonstrate that even relatively low levels of fetal paracetamol exposure are associated with substantial alterations in ovarian morphology and activity. Exposed girls exhibited approximately 50% reductions in ovarian volume and follicle number. In addition, the uterine volume and breast tissue diameter were reduced, indicating compromised estradiol production. Associations were consistent and dose dependent across complementary exposure assessment strategies, and the principal results were replicated in an independent cohort followed into adolescence, suggesting that the observed alterations may persist beyond early life.

These human data align closely with a substantial body of experimental evidence. Multiple independent animal studies have consistently demonstrated reduced ovarian follicle numbers following prenatal paracetamol exposure(Aleixo *et al*., 2020; Dean *et al*., 2016; Holm *et al*., 2016; Johansson *et al*., 2016; Rossitto *et al*., 2019; Wu *et al*., 2024), providing biological plausibility and supporting a potential causal interpretation of the associations observed in the present study.

Our observations align with current understanding of fetal ovarian biology, and several biological mechanisms have been proposed that could explain the associations. Paracetamol is suggested to inhibit the intense mitosis of germ cells, thereby reducing the number of oogonia entering the first meiotic division (Dean *et al*., 2016; Holm *et al*., 2016; Hurtado-Gonzalez *et al*., 2018; Rossitto *et al*., 2019). This effect may occur through direct inhibition of cell proliferation, potentially via targeting ribonucleotide reductase and subsequent blockade of DNA synthesis (Kristensen *et al*., 2024). In addition, paracetamol inhibits COX enzyme activity, leading to reduced prostaglandin synthesis and decreased expression of key pluripotency and anti-apoptotic markers in germ cells (Bayne *et al*., 2022; Dean *et al*., 2016; Hurtado-Gonzalez *et al*., 2018). Moreover, reduced prostaglandin levels delay the critical transition from germ cell mitosis to meiosis resulting in intensified germ cell apoptosis, further diminishing the final pool of primordial follicles (Dean *et al*., 2016; Holm *et al*., 2016; Martins da Silva *et al*., 2004; Rossitto *et al*., 2019).

The functional establishment of the female HPG axis during minipuberty provides a unique window to assess ovarian activity (Fischer, Mola, Rom, *et al*., 2024; Ljubicic *et al*., 2022). AMH is produced exclusively by granulosa cells surrounding small antral follicles, and in adult women, AMH levels and the antral follicle count serve as non-invasive proxies for the size of the ovarian reserve of primordial follicles (Andersen *et al*., 2010; Gougeon, 1996; Hansen *et al*., 2011; Jeppesen *et al*., 2013; Mamsen *et al*., 2021). In adult women, age specific AMH is therefore related to age at menopause (Depmann *et al*., 2018). During childhood, AMH (SDS) is relatively stable from infancy to adolescence (Hagen *et al*., 2023). In the present study, AMH levels were reduced in infant girls exposed exclusively in early fetal, suggesting that fetal paracetamol exposure may diminish the ovarian reserve. Whether these differences in early postnatal life carry long-term clinical significance, including potential implications for reproductive lifespan, remains to be further elucidated and requires confirmation in future studies with extended follow-up. In addition, exposure was associated with reduced sizes of estrogen responsive tissues. At minipuberty, uterine volume and diameter of breast tissue correlate with circulating estradiol levels (Fischer, Mola, Rom, *et al*., 2024; Kuiri-Hänninen *et al*., 2013), and although a single estradiol concentration at a single evaluation in the present study was not associated with exposure, we speculate, that cumulative lower estradiol secretion from fewer ovarian follicles contribute to reduced growth of uterus and breast tissue.

The present study is the first human cohort specifically designed to assess associations between prenatal exposure to paracetamol and ovarian development and exposure assessment is a critical strength of this study. We combined frequently maternal self-reported data with urinary paracetamol measurements, providing both subjective and objective exposure indices. The strong dose-response trends, particularly for ovarian and uterine volume, reinforce the robustness of the findings. Paracetamol was detected in all urine samples, reflecting widespread environmental exposure likely originating from aniline metabolism (Nielsen *et al*., 2015; Pot E *et al*., 2022). As no standardized cutoff exists for recent intake, percentile-based thresholds were applied, which yielded consistent results with reported use and suggest biological relevance even at low exposure levels.

Unique to this study is the thorough assessment of ovarian activity in infancy including detailed ovarian and uterine morphology by transabdominal ultrasound scans, the most appropriate method for evaluating internal genitalia morphology in prepubertal girls.

A key strength of this study is replication of the results in an independent confirmatory cohort. Although this cohort was not primarily designed to assess maternal paracetamol use and exposure therefore is most likely highly underreported, similar associations with ovarian and uterine morphology were observed during puberty and adolescence. These observations support the associations seen in the COPANA cohort and suggest the possibility that the observed differences may persist beyond infancy, but the clinical relevance warrant further investigation.

Hormonal assessments during adolescence did not show clear associations with exposure, likely due to menstrual cycle variability. Limited exposure data from mid-to-late pregnancy in the confirmatory cohort may have further restricted the detection of associations with follicle numbers.

Due to the study design, residual confounding by indication cannot be completely excluded, although results were robust after accounting for fever and other maternal factors. The study population was predominantly Caucasian and term-born, which may limit generalizability. The analytic design of the current study limited our ability to evaluate whether frequency or patterns of paracetamol use influenced the observed associations; however, even with a dichotomous yes/no exposure definition across two broad exposure windows, we observed consistent and biologically plausible associations across cohorts and developmental stages.

Paracetamol is available over the counter and is widely used during pregnancy (Ersbøll *et al*., 2015; Lind *et al*., 2017; Taagaard *et al*., 2023). Although the present findings show that fetal exposure was associated with ovarian morphology and activity in infancy, the extent to which these early-life observations translate into long-term effects on reproductive function remains to be further evaluated.

While certain clinical situations, such as severe maternal fever, require paracetamol use(Antoun *et al*., 2021; Dreier *et al*., 2014) current guidelines recommend minimal dosage and duration of therapy (European Medicines Agency, 2024). Exposure levels in this study fall within these recommendations, raising concern that current guidance may not be sufficient. Non-pharmacological alternatives and enhanced education for pregnant women may help reduce unnecessary exposure.

## 7. Conclusion

Mild to moderate doses of prenatal paracetamol exposure was associated with ovarian morphology and activity in infant girls, including reduced ovarian size, fewer follicle, and altered reproductive hormone levels, as well as smaller uterine volume and diameter of breast tissue. These associations were supported by an independent confirmatory cohort, suggesting persistent effects in adolescence.

## Author contributions

MBF and CH had full access to the study data and take responsibility for the integrity of the data and the accuracy of the data analysis. The corresponding author attests that all listed authors meet authorship criteria and that no others meeting the criteria have been omitted. The lead author (the manuscript’s guarantor) affirms that the manuscript is an honest, accurate, and transparent account of the study being reported to that no important aspects of the study have been omitted to and that any discrepancies from the study as planned (and, if relevant, registered) have been explained. MBF and CH designed the COPANA cohort and applied for grants. MBF and GM coordinated the study and examined the girls. MBF did the statistical analyses. MBF, CH and JHP interpreted the data. MBF drafted the original manuscript, which was reviewed and edited by all other authors. All authors approved the final submitted version.

KMM designed and conducted the independent mother child cohort study, MA performed all clinical examinations during adolescence.

## Acknowledgements

The participation of the families is gratefully acknowledged. We further thank the laboratory technicians at the Hormone-, Molecular- as well as the Chemistry Laboratory at the Department of Growth and Reproduction.

The funders had no role in considering the study design or in the collection, analysis, interpretation of data, writing of the report, or decision to submit the article for publication. This research was supported by Rigshospitalets Research Council under grant (E-22717-21), Læge Sofus Carl Emil Friis og hustru Doris Friis’ Legat (F-23936-01), Aase og Ejnar Danielsens Foundation (20-10-0367), Helsefonden (20-B-0388), Axel Muusfeldt Foundation (2020-0385) and The Danish Centre for Endocrine Disrupting Substances (CeHoS) (2022-23219). The authors have nothing to declare.

## Data sharing statement

All data will be made available from the corresponding author at on reasonable request and according to GDPR rules.

## Supplementary material

**Supplementary Table S1.** Hormone analyses in infant blood samples (n=269). Listed by priority of analyses.

|  | Method/Assay | Manufacturer | N* | Samples above<br>LOD (%) | LOD |
| --- | --- | --- | --- | --- | --- |
| Anti-Müllerian hormone (AMH) | Chemiluminescence immunoassay | Access2, Beckman Coulter, Brea, CA, USA | 264 | 100 | 0.14 pmol/L |
| Follicle-stimulating hormone (FSH) | Chemiluminescence immunoassay | Atellica, Siemens NY, Healthineers, Tarrytown, USA | 264 | 100 | 0.3 IU/L |
| Luteinizing hormone (LH) | Chemiluminescence immunoassay | Atellica, Siemens Healthineers, Tarrytown, NY, USA | 264 | 47 | 0.07 IU/L |
| Inhibin B | Enzyme-linked immunosorbent assay | Immunotech, Beckman Coulter, Brea, CA, USA | 265 | 97 | 3 ng/L |
| Estradiol | LC/MS-MS | *In-house* | 265 | 89 | 4.0 pmol/L |
| Progesterone | LC/MS-MS | *In-house* | 269 | 100 | 0.036 nmol/L |
| Androstenedione | LC/MS-MS | *In-house* | 269 | 100 | 0.031 nmol/L |
| Testosterone | LC/MS-MS | *In-house* | 269 | 100 | 0.031 nmol/L |

LOD: limit of detection, LC/MS-MS: liquid chromatography-tandem mass spectrometry. \*Because the analyses require varying amounts of sample volume, the number of completed tests does not always align with the prioritization.

**Supplementary Table S2.** Adjusted and unadjusted associations between maternal self-reported use of paracetamol and predefined primary and secondary outcomes in girls exposed exclusively during early fetal life (GA < 17 weeks, n = 22) compared to unexposed girls (n = 143).

|  | Unadjusted | Adjusted |
| --- | --- | --- |
|  | Estimate (95% CI) | Estimate (95% CI) |
| Uterine volume (cm<sup>3</sup>) | **-0.19 (-0.39 ; 0.01)\*** | **-0.2 (-0.43 ; 0.03)\*** |
| Endometrial thickness (mm) | -0.1 (-0.35 ; 0.16) | 0 (-0.03 ; 0.03) |
| Ovarian volume (cm<sup>3</sup>) | **-0.12 (-0.22 ; -0.02)\*\*** | **-0.1 (-0.19 ; -0.01)\*\*** |
| Sum of total follicles | -0.09 (-1.32 ; 1.13) | 0.15 (-1.1 ; 1.4) |
| Breast tissue diameter (cm) | 0.02 (-0.21 ; 0.24) | -0.04 (-0.26 ; 0.18) |
| Fetal AGD (cm) | 0.02 (-0.09 ; 0.14) | 0 (-0.11 ; 0.11) |
| AGDac (cm) | 0.1 (-0.1 ; 0.31) | -0.01 (-0.17 ; 0.15) |
| AGDaf (cm) | 0.02 (-0.15 ; 0.2) | 0.06 (-0.14 ; 0.26) |
| AMH (SDS) | **-0.41 (-0.83 ; 0)\*\*** | **-0.45 (-0.87 ; -0.03)\*\*** |
| Inhibin B (SDS) | 0.11 (-0.35 ; 0.58) | 0.13 (-0.33 ; 0.59) |
| FSH (SDS) | 0.33 (-0.1 ; 0.75) | **0.39 (-0.07 ; 0.85)\*** |
| Estradiol (SDS) | -0.27 (-0.76 ; 0.22) | -0.24 (-0.72 ; 0.24) |

Estimated mean differences with 95% confidence intervals (CI) from linear regression models, adjusted and unadjusted. Model adjusted for maternal polycystic ovarian syndrome status, BMI before pregnancy, alcohol and nicotine use in the first trimester, conception by in vitro fertilization, maternal use of acetylic salicylic acid and other non-steroidal inflammatory drugs, infant paracetamol treatment and infant age at examination (uterine volume, AGDaf, AGDac, breast tissue diameter). Fetal AGD is moreover adjusted for GA at time of ultrasound scan. SDS: standard deviation score, AGD: anogenital distance, AGDaf: anogenital distance (ano-fourchettal), AGDac: anogenital distance (ano-clitoral), FSH: follicle stimulating hormone, AMH: anti-Müllerian hormone. \*indicate associations with a p-value $0.05 \leq 0.1$ and \*\*indicate a p-value $\leq 0.05$ .

**Supplementary Table S3.** Multiple regression analyses estimating the independent association of each exposure period between fetal exposure to paracetamol (yes/no) in girls exposed during early fetal life (GA < 17 weeks, n = 92) and/or during mid-late fetal life (GA ≥ 17 weeks, n = 137).

|  | Estimate (95% CI) |
| --- | --- |
| **Fetal AGD (cm)** |  |
| Paracetamol in early fetal life (GA < 17 weeks) | -0.015 (-0.08 ; 0.05) |
| Paracetamol in mid-late fetal life (GA ≥ 17 weeks) | 0.008 (-0.052 ; 0.068) |
| **Uterine volume (cm<sup>3</sup>)** |  |
| Paracetamol in early fetal life (GA < 17 weeks) | **-0.141 (-0.26 ; -0.022)\*\*** |
| Paracetamol in mid-late fetal life (GA ≥ 17 weeks) | -0.018 (-0.145 ; 0.109) |
| **Endometrial thickness (mm)** |  |
| Paracetamol in early fetal life (GA < 17 weeks) | -0.039 (-0.2 ; 0.122) |
| Paracetamol in mid-late fetal life (GA ≥ 17 weeks) | 0.032 (-0.127 ; 0.191) |
| **Ovarian volume (cm<sup>3</sup>)** |  |
| Paracetamol in early fetal life (GA < 17 weeks) | -0.042 (-0.144 ; 0.06) |
| Paracetamol in mid-late fetal life (GA ≥ 17 weeks) | 0.003 (-0.105 ; 0.111) |
| **Sum of total follicles** |  |
| Paracetamol in early fetal life (GA < 17 weeks) | **0.621 (-0.027 ; 1.269)\*** |
| Paracetamol in mid-late fetal life (GA ≥ 17 weeks) | **-0.735 (-1.332 ; -0.138)\*\*** |
| **AGDaf (cm)** |  |
| Paracetamol in early fetal life (GA < 17 weeks) | 0.046 (-0.047 ; 0.139) |
| Paracetamol in mid-late fetal life (GA ≥ 17 weeks) | 0.02 (-0.066 ; 0.106) |
| **AGDac (cm)** |  |
| Paracetamol in early fetal life (GA < 17 weeks) | 0.087 (-0.042 ; 0.216) |
| Paracetamol in mid-late fetal life (GA ≥ 17 weeks) | **-0.119 (-0.234 ; -0.004)\*\*** |
| **Breast tissue diameter (cm)** |  |
| Paracetamol in early fetal life (GA < 17 weeks) | 0.049 (-0.09 ; 0.188) |
| Paracetamol in mid-late fetal life (GA ≥ 17 weeks) | -0.068 (-0.196 ; 0.06) |
| **AMH (SDS)** |  |
| Paracetamol in early fetal life (GA < 17 weeks) | 0.059 (-0.235 ; 0.353) |
| Paracetamol in mid-late fetal life (GA ≥ 17 weeks) | 0.12 (-0.143 ; 0.383) |
| **Inhibin B (SDS)** |  |
| Paracetamol in early fetal life (GA < 17 weeks) | **0.225 (-0.024 ; 0.474)\*** |
| Paracetamol in mid-late fetal life (GA ≥ 17 weeks) | -0.097 (-0.333 ; 0.139) |
| **FSH (SDS)** |  |
| Paracetamol in early fetal life (GA < 17 weeks) | 0.004 (-0.246 ; 0.254) |
| Paracetamol in mid-late fetal life (GA ≥ 17 weeks) | -0.075 (-0.319 ; 0.169) |
| **Estradiol (SDS)** |  |
| Paracetamol in early fetal life (GA < 17 weeks) | 0.02 (-0.273 ; 0.313) |
| Paracetamol in mid-late fetal life (GA ≥ 17 weeks) | -0.084 (-0.364 ; 0.196) |

Estimated mean differences with 95% confidence intervals (CI) from linear regression models. Model adjusted for maternal polycystic ovarian syndrome status, BMI before pregnancy, alcohol and nicotine use in the first trimester, conception by in vitro fertilization, maternal use of acetylic salicylic acid and other non-steroidal anti-inflammatory drugs, infant paracetamol treatment and infant age at examination (uterine volume, AGDaf, AGDac, breast tissue diameter). SDS: standard deviation score, FSH: follicle stimulating hormone, AGDaf: anogenital distance (ano-fourchettal), AGDac: anogenital distance (ano-clitoral), AMH: anti-Müllerian hormone. \*indicate associations with a p-value of 0.05 ≤ 0.1 and \*\*indicate a p-value ≤ 0.05.

**Supplementary Table S4.** Percentile distribution of accumulated maternal self-reported doses of paracetamol use during pregnancy (n = 302) as well as first trimester urinary paracetamol (U-paracetamol) concentrations (n = 299).

| Percentile | Dose of paracetamol |  | U-paracetamol |  |
| --- | --- | --- | --- | --- |
|  | mg | n | ng/mL | n |
| 50 | 750.0 | 151/151 | 64.4 | 149/150 |
| 75 | 3375.0 | 226/76 | 134.9 | 224/75 |
| 95 | 20000.0 | 285/17 | 3008.6 | 284/15 |

**Supplementary Table S5.** Associations between maternal self-reported paracetamol use and predefined primary and secondary outcomes in girls exposed during early fetal life (GA < 17 weeks, n = 92) or during mid-late fetal life (GA ≥ 17 weeks, n = 67), compared to unexposed girls (n = 143). All analyses exclude infants whose mothers reported fever as an indication for paracetamol use.

|  | Early fetal life<br>Estimate (95% CI) | Mid-Late fetal life<br>Estimate (95% CI) |
| --- | --- | --- |
| Uterine volume (cm<sup>3</sup>) | -0.12 (-0.29 ; 0.05) | -0.13 (-0.4 ; 0.14) |
| Endometrial thickness (mm) | -0.04 (-0.24 ; 0.15) | -0.07 (-0.38 ; 0.24) |
| Ovarian volume (cm<sup>3</sup>) | -0.05 (-0.15 ; 0.06) | -0.07 (-0.27 ; 0.13) |
| Sum of total follicles | -0.05 (-0.15 ; 0.06) | **-1.66 (-2.96 ; -0.35)\*\*** |
| Breast tissue diameter (cm) | 0.06 (-0.08 ; 0.2) | -0.09 (-0.29 ; 0.11) |
| Fetal AGD (cm) | 0.01 (-0.05 ; 0.07) | 0 (-0.07 ; 0.07) |
| AGDaf (cm) | 0.05 (-0.04 ; 0.14) | -0.01 (-0.15 ; 0.12) |
| AGDac (cm) | 0.03 (-0.11 ; 0.17) | -0.12 (-0.31 ; 0.07) |
| AMH (SDS) | 0.15 (-0.17 ; 0.47) | -0.09 (-0.54 ; 0.36) |
| Inhibin B (SDS) | 0.07 (-0.19 ; 0.34) | -0.27 (-0.66 ; 0.12) |
| FSH (SDS) | -0.04 (-0.31 ; 0.23) | 0 (-0.42 ; 0.42) |
| Estradiol (SDS) | -0.08 (-0.39 ; 0.22) | -0.32 (-0.78 ; 0.13) |

Estimated mean differences with 95% confidence intervals (CI) from linear regression models. SDS: standard deviation score, AGD: anogenital distance, AGDaf: anogenital distance (ano-fourchette), AGDac: anogenital distance (ano-clitoral), FSH: follicle stimulating hormone, AMH: anti-Müllerian hormone. \*indicate associations with a p-value of $0.05 \leq 0.1$ and \*\*indicate a p-value $\leq 0.05$ .

**Supplementary Table S6.** Maternal and infant characteristics stratified by exposure to paracetamol and/or NSAIDs in fetal life (n = 179) compared to unexposed controls (n=1,012) from the Copenhagen mother-child cohort.

|  | Total |  | Unexposed |  | Exposed |  |
| --- | --- | --- | --- | --- | --- | --- |
|  | N | Mean (±SD) / n (%) | N | Mean (±SD) / n (%) | N | Mean (±SD) / n (%) |
| Gestational age at birth (weeks) | 1142 | 39.63 (1.95) | 964 | 39.63 (1.95) | 178 | 39.64 (1.95) |
| Maternal thyroid disease or diabetes | 1191 | 24 (2) | 1012 | 19 (2) | 179 | 5 (3) |
| Maternal smoking | 1077 | 260 (24) | 906 | 230 (25) | 171 | **30 (18)\*\*** |
| Birth weight (kg) | 879 | 3.43 (0.55) | 734 | 3.41 (0.53) | 145 | **3.52 (0.61)\*\*** |
| Birth length (cm) | 876 | 50.57 (2.24) | 731 | 50.51 (2.22) | 145 | **50.90 (2.32)\*** |
| **Infancy** | **1083** |  | **904** |  | **179** |  |
| Age (days) | 1076 | 101.11 (24.01) | 897 | 101.41 (24.14) | 179 | 99.61 (23.35) |
| BMI (kg/m<sup>2</sup>) | 1069 | 16.24 (1.46) | 891 | 16.22 (1.46) | 178 | 16.33 (1.45) |
| **Puberty** | **563** |  | **456** |  | **107** |  |
| Age (years) | 563 | 10.72 (1.56) | 456 | 10.72 (1.65) | 107 | 10.71 (1.14) |
| BMI (kg/m<sup>2</sup>) | 533 | 17.37 (2.59) | 432 | 17.29 (2.51) | 101 | 17.71 (2.91) |
| Tanner breast stage 1 | 118 | 15 (13) | 96 | 13 (14) | 22 | 2 (9.1) |
| Tanner breast stage 2 | 118 | 31 (27) | 96 | 25 (26) | 22 | 6 (27.3) |
| Tanner breast stage 3 | 118 | 34 (29) | 96 | 24 (25) | 22 | **10 (45.5)\*** |
| Tanner breast stage 4 | 118 | 36 (31) | 96 | 32 (33) | 22 | 4 (18.2) |
| Tanner breast stage 5 | 118 | 2 (1.7) | 96 | 2 (2) | 22 | 0 (0.0) |
| **Adolescence** | **317** |  | **266** |  | **51** |  |
| Age (years) | 317 | 16.05 (0.89) | 266 | 16.10 (0.93) | 51 | **15.78 (0.59)\*\*** |
| BMI (kg/m<sup>2</sup>) | 314 | 21.52 (3.04) | 263 | 21.54 (3.10) | 51 | 21.46 (2.74) |

AMH: anti-Müllerian hormone, FSH: follicle stimulating hormone, LH: luteinizing hormone. P-values for continuous variables were obtained from a Student's t-test. Girls using hormonal contraceptives are removed from adolescent analyses.\*indicate associations with a p-value of 0.05 ≤ p < 0.1 and \*\*indicate a p-value ≤ 0.05.

**Supplementary Table S7.** Summaries of outcomes in girls exposed during pregnancy (n = 179) compared to unexposed girls (n = 1,012) from the Copenhagen mother-child cohort.

|  | Total |  | Unexposed |  | Exposed |  |
| --- | --- | --- | --- | --- | --- | --- |
|  | N | Mean (±SD) | N | Mean (±SD) | N | Mean (±SD) |
| **Infancy** | **1083** |  | **904** |  | **179** |  |
| AMH (pmol/L) | 442 | 17.18 (14.25) | 370 | 16.82 (13.23) | 72 | 19.06 (18.65) |
| Inhibin B (pg/ml) | 552 | 81.86 (51.24) | 466 | 83.15 (51.53) | 86 | 74.86 (49.33) |
| FSH (IU/L) | 587 | 4.99 (4.25) | 496 | 4.97 (4.03) | 91 | 5.12 (5.29) |
| **Puberty** | **563** |  | **456** |  | **107** |  |
| AMH (pmol/L) | 399 | 23.36 (14.62) | 320 | 23.64 (14.83) | 79 | 22.25 (13.77) |
| Inhibin B (pg/ml) | 398 | 42.42 (28.16) | 319 | 43.37 (28.75) | 79 | 38.59 (25.41) |
| FSH (IU/L) | 398 | 3.29 (1.94) | 319 | 3.30 (1.97) | 79 | 3.27 (1.81) |
| LH (IU/L) | 398 | 1.61 (2.64) | 319 | 1.68 (2.82) | 79 | 1.31 (1.68) |
| MRI: Uterine volume (cm<sup>3</sup>) | 61 | 18.16 (15.10) | 53 | 19.47 (15.40) | 8 | **9.46 (9.63)\*\*** |
| MRI: Ovarian volume (cm<sup>3</sup>) | 84 | 6.91 (3.81) | 67 | 7.05 (4.02) | 17 | 6.40 (2.93) |
| MRI: Sum of total follicles | 87 | 20.99 (10.51) | 70 | 21.10 (10.81) | 17 | 20.53 (9.45) |
| **Adolescence** | **317** |  | **266** |  | **51** |  |
| AMH (pmol/L) | 305 | 28.05 (16.54) | 253 | 28.16 (16.77) | 52 | 27.52 (15.49) |
| Inhibin B (pg/ml) | 294 | 66.96 (37.03) | 245 | 67.49 (37.72) | 49 | 64.29 (33.63) |
| FSH (IU/L) | 294 | 4.91 (2.57) | 245 | 4.81 (2.62) | 49 | 5.38 (2.28) |
| LH (IU/L) | 294 | 4.10 (5.80) | 245 | 3.90 (5.53) | 49 | 5.12 (6.97) |
| TAUS: Uterine volume (cm<sup>3</sup>) | 288 | 46.87 (12.44) | 240 | 47.03 (12.57) | 48 | 46.06 (11.85) |
| TAUS: Ovarian volume (cm<sup>3</sup>) | 288 | 15.92 (11.32) | 239 | 16.30 (12.27) | 49 | **14.07 (4.09)\*\*** |
| TAUS: Sum of total follicles | 287 | 19.17 (4.37) | 239 | 19.12 (4.32) | 48 | 19.46 (4.64) |

AMH: anti-Müllerian hormone, FSH: follicle stimulating hormone, LH: luteinizing hormone, MRI: Magnetic Resonance Imaging, TAUS: transabdominal ultrasound scan. P values for continuous variables were obtained from a Student's t-test. Girls using hormonal contraceptives are removed from adolescent analyses. \*\*indicate a p-value ≤ 0.05.

**Supplementary Table S8.** Adjusted and unadjusted associations between maternal self-reported use of paracetamol and/or non-steroidal anti-inflammatory drugs in girls exposed during pregnancy (n = 179) compared to unexposed girls (n = 1,012) from the Copenhagen mother-child cohort.

|  | Unadjusted | Adjusted |
| --- | --- | --- |
|  | Estimate (95% CI) | Estimate (95% CI) |
| **Infancy** |  |  |
| AMH (pmol/L) | 2.24 (-2.25 ; 6.72) | 2.29 (-2.11 ; 6.69) |
| Inhibin B (pg/ml) | -8.29 (-19.66 ; 3.08) | -8.03 (-19.21 ; 3.15) |
| FSH (IU/L) | 0.16 (-0.98 ; 1.29) | 0.16 (-0.98 ; 1.30) |
| **Puberty** |  |  |
| AMH (pmol/L) | -1.38 (-4.81 ; 2.04) | 1.97 (-4.08 ; 8.03) |
| Inhibin B (pg/ml) | -4.77 (-11.17 ; 1.63) | **-14.12 (-25.20 ; -3.03)\*\*** |
| FSH (IU/L) | -0.03 (-0.49 ; 0.42) | **-0.61 (-1.21 ; -0.00)\*\*** |
| LH (IU/L) | -0.37 (-0.85 ; 0.11) | -0.37 (-0.88 ; 0.14) |
| MRI: Uterine volume (cm<sup>3</sup>) | **-10.01 (-17.49 ; -2.54)\*\*** | **-4.11 (-7.29 ; -0.92)\*\*** |
| MRI: Ovarian volume (cm<sup>3</sup>) | -0.64 (-2.30 ; 1.01) | -0.05 (-1.33 ; 1.23) |
| MRI: Sum of total follicles | -0.57 (-5.60 ; 4.47) | -0.36 (-5.21 ; 4.50) |
| **Adolescence** |  |  |
| AMH (pmol/L) | -0.64 (-5.29 ; 4.01) | -1.19 (-6.42 ; 4.03) |
| Inhibin B (pg/ml) | -3.20 (-13.65 ; 7.24) | -5.69 (-17.06 ; 5.67) |
| FSH (IU/L) | 0.57 (-0.14 ; 1.28) | 0.46 (-0.28 ; 1.21) |
| LH (IU/L) | 1.22 (-0.84 ; 3.27) | 1.12 (-1.08 ; 3.32) |
| TAUS: Uterine volume (cm<sup>3</sup>) | -0.97 (-4.65 ; 2.71) | -0.59 (-4.40 ; 3.22) |
| TAUS: Ovarian volume (cm<sup>3</sup>) | **-2.23 (-4.15 ; -0.31)\*\*** | **-2.76 (-4.82 ; -0.70)\*\*** |
| TAUS: Sum of total follicles | 0.34 (-1.07 ; 1.75) | 0.17 (-1.31 ; 1.65) |

Estimated mean differences with 95% confidence intervals (CI) from linear regression models.
Model adjusted for maternal thyroid disease or diabetes and gestational week at birth. Further, pubertal outcomes are adjusted for Tanner stage at time of examination and girls using hormonal contraceptives are removed from adolescent analyses. AMH: anti-Müllerian hormone, FSH: follicle stimulating hormone, LH: luteinizing hormone, MRI: Magnetic Resonance Imaging, TAUS: transabdominal ultrasound scan. \*\*indicate associations with a p-value ≤ 0.05.

**Supplementary Fig. S1.**
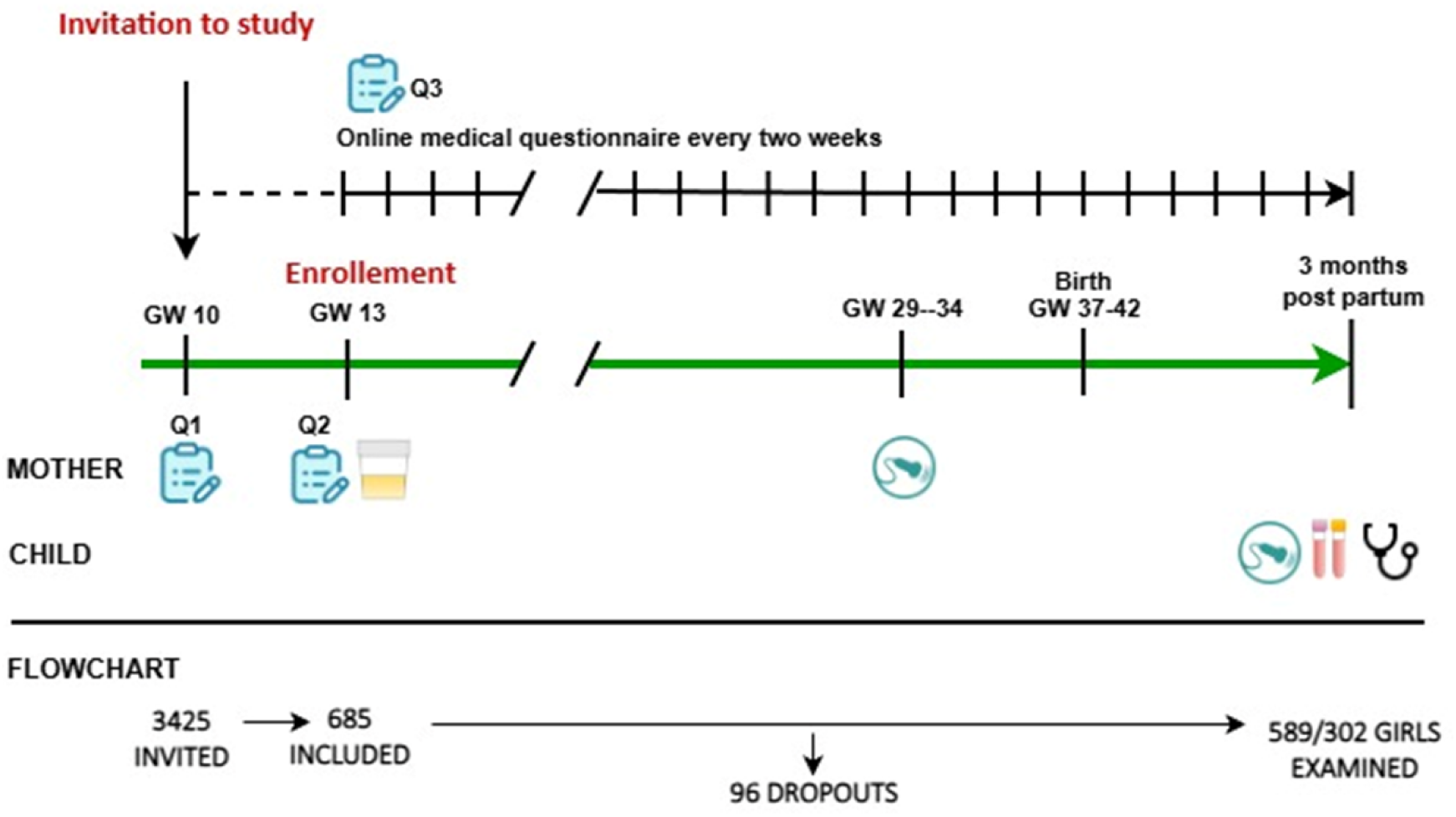
Study timeline showing study visits, questionnaires and sample collection GW: gestational week Q1: Routine web-based questionnaire Q2: Study specific questionnaire on general- and reproductive health Q3: Study specific questionnaire on medical use every two weeks

